# Characteristics and outcomes of Acute Respiratory Distress Syndrome related to COVID-19 in Belgian and French Intensive Care Units according to antiviral strategies. The COVADIS multicenter observational study

**DOI:** 10.1101/2020.06.28.20141911

**Authors:** David Grimaldi, Nadia Aissaoui, Gauthier Blonz, Giuseppe Carbutti, Romain Courcelle, Stephane Gaudry, Alain D’hondt, Julien Higny, Geoffrey Horlait, Sami Hraiech, Laurent Lefebvre, Francois Lejeune, Andre Ly, Michael Piagnerelli, Bertrand Sauneuf, Nicolas Serck, Thibaud Soumagne, Piotr Szychowiak, Julien Textoris, Benoit Vandenbunder, Christophe Vinsonneau, Jean-Baptiste Lascarrou, for the COVADIS study group

## Abstract

**Background:** Limited data are available for antiviral therapy efficacy especially for the most severe patients under mechanical ventilation suffering from Covid-19 related Acute Respiratory Distress Syndrome (ARDS).

**Methods:** Observational multicenter cohort of patients with moderate to severe Covid-19 ARDS, comparing antiviral strategies (none, hydroxychloroquine (HCQ), lopinavir/ritonavir (L/R), others (combination or remdesivir). The primary end-point was the day-28 ventilator free days (VFD), patients which died before d28 were considered as having 0 VFD. The variable was dichotomized in patients still ventilated or dead at day 28 vs patients being extubated and alive at day 28 (VFD = or >0).

**Results:** We analyzed 376 patients (80 with standard of care (SOC), 49 treated with L/R, 197 with HCQ, and 50 others). The median number of d28-VFD was 0 (IQR 0-13) and was different across the different groups (P=0.01), the SOC patients having the highest d28-VFD. A multivariate logistic regression including antiviral strategies, showed that age (OR 0.95 CI95%:0.93-0.98), male gender (OR 0.53 CI95%:0.31-0.93), Charlson score (OR 0.85 CI95%:0.73-0.99) and plateau pressure (OR 0.94 CI95%:0.88-0.99) were associated with having 0 d28-VFD whereas P/F ratio (OR 1.005 CI95%:1.001-1.010) was associated with having ≥1 d28-VFD (ie. being extubated and alive). Acute kidney injury (AKI) was frequent (64%), its incidence was different across the patients’ groups (P=0.01). In a post-hoc logistic multivariate regression apart from demographics characteristics and comorbidities, the use of L/R (administered to 81 of 376 patients was associated with occurrence of AKI (OR 2.07 CI95%:1.17-3.66) and need for renal replacement therapy (RRT).

**Conclusion:** In this observational study of moderate to severe Covid-19 ARDS patients, we did not observed a benefit of treating patients with any specific antiviral treatment. We observed an association between L/R treatment and occurrence of AKI and need for RRT.

**Take home message:** Any specific COVID-19 antiviral treatment is associated with higher ventilator free days at day 28 as compared to no antiviral treatment for patient in ICU under invasive mechanical ventilation. Lopinavir/ritonavir is associated with an increased risk of acute kidney injury.

**Tweet:** COVID-19: Insights from ARDS cohort: no signal of efficacy for antiviral treatments. Lopinavir/ritonavir may be associated with AKI and need for RRT.

## Introduction

Since December 2019, Coronavirus Disease 2019 (COVID-19) related to infection by severe acute respiratory syndrome coronavirus 2 (SARS-CoV-2) emerged in Wuhan, China, and rapidly spread throughout China, Asia and the world [1]. COVID-19 can have different clinical presentations but respiratory symptoms predominate especially in patients admitted to intensive care units (ICU) [2]. The respiratory disease appears to be a very homogenous entity [3]. It mainly affects people older than 50 years, predominantly men with cardiovascular comorbidities [2]. It is characterized by severe hypoxemia, radiological ground glass opacities and especially crazy paving. Its evolution is prolonged with an aggravation phase 7-10 days after symptoms onset [4] leading to death between 3% for patients in the ward [5] to 60% of patients in the ICU [6].

Although many unknowns persist, such a great homogeneity of presentation was previously unseen in acute pathologies leading to intensive care unit (ICU). Despite expert recommendations that were implemented quickly [7], management was not based on high levels of evidence during the West-European first wave (March-April 2020). The treatments applied vary from one country to another, from one center to another and even from one patient to another. A desperate search for efficient antiviral treatment has been ongoing since the epidemic started. First candidates were already-developed molecules that demonstrated *in vitro* effect, mainly remdesivir [8], lopinavir/ritonavir [7] and hydroxychloroquine [9]. The last two are already on the market with an acceptable safety profile; however, they have not been used widely in critically ill patients so that potential previously unknown side effects may exist. Even some randomized clinical trials are now available such as for Remdesivir [10], or for lopinavir/ritonavir alone [7] or in association [11], only a small proportion of patients included were under mechanical ventilation not allowing efficacy and safety analysis for patients sicker than in conventional ward.

The main objective of this study was to set up an observational study of patients suffering from moderate to severe COVID-19 related ARDS, by collecting the strategies used in Belgian and French ICUs, in order to be able to detect a possible signal of efficacy or deleterious effects of used therapies. We reported here the data regarding the antiviral strategies.

### Patients and methods

This study was compliant with STROBE guidelines [12].

### Study design and setting

The COVADIS project is observational and regroups 21 ICUs in France (n=12) and Belgium (n=9).

### Patient selection

Inclusion criteria were:

- Age older than 18 years,
- moderate to severe ARDS according to Berlin definition [13] (PaO2/FiO2 ratio < 200mmHg with a PEEP of at least 5mmHg receiving invasive ventilation),
- positive SARS-CoV-2 reverse transcriptase polymerase chain reaction (PCR) regardless of site sampled (patient with negative PCR but chest CT scan with abnormalities such as crazy paving were not included).

Non-inclusion criteria were:

- Cardiac arrest before intensive care unit admission,
- Extra Corporeal Mobile Oxygenation (ECMO) requirement within first 24hrs of ICU length,
- Chronic Obstructive Pulmonary Disease with Gold class 3 or 4 [14], or home oxygen.

### Data collection

For this observational multicenter study, all consecutive COVID-19 patients were screened in the participating centers. Patients fulfilling inclusion and non-inclusion criteria were included in participating ICUs between March 10, 2020 and April 12, 2020. Each local investigator filled an eCRF to collect data (Castor EDC, Amsterdam, The Netherlands). We recorded demographics data, known medical history and co-morbidities using the Charlson score [15], with the addition of history of chronic hypertension. We collected data on management interventions delivered during hospitalization including settings of MV after intubation, duration of MV, administration of advanced therapies for acute respiratory failure (neuromuscular blocking agents, inhaled pulmonary vasodilators, prone-positioning, and extracorporeal membrane oxygenation), anti-viral treatment and immunomodulatory agents (interleukin-6-receptor antagonists and corticosteroids) with time from onset of symptoms to initiation and occurrence of acute kidney injury (AKI), acute cardiac injury (defined as a rise in troponin level over 10 times the normal threshold or the needs for inotrope), pulmonary embolism and deep venous thrombosis.

### Primary outcomes

Considering the absence of efficacy or safety data on antiviral treatments in the area of critically ill patients requiring invasive mechanical ventilation, we decided to choose a composite outcome which included death and length of mechanical ventilation: number of ventilator free days (VFD) at day 28 [16]. VFD at day 28 was determined as follow:

- VFDs□= □ 0 if subject dies within 28 days of mechanical ventilation,
- VFDs□= □ 28□− □ *x* if successfully liberated from ventilation *x* days after initiation,
- VFDs□= □ 0 if the subject is mechanically ventilated for >28 days.

The variable was dichotomized in patients still ventilated or dead at day 28 vs patients being extubated and alive at day 28 (VFD = or >0).

### Secondary outcomes

- Day 14 and Day 28 survival
- Ventilator mode at day 14 according to pre-defined 4 categories: patient under Volume/Pressure Assisted Controlled or ECMO, Pressure Support mode, Spontaneous breathing while extubated, Death
- Need for ECMO after the first 24h
- Cardiac dysfunction (defined as plasmatic Troponin level upper than 10 time upper normal range or need for inotrope (dobutamine, epinephrine, milrinone, and/or levosimendan),
- AKI defined by a rise in serum creatinine of at least 50% as defined in KDIGO stage 1 [17], and classified as none, present without need for renal replacement therapy (RRT), present with need for RRT,
- Deep venous thrombosis, and pulmonary embolism.

### Statistical analysis

As many candidate for specific antiviral treatment are currently under evaluation, our variable of interest was use of anti-viral treatment according to one of the pre-specified following category: none (Standard of care), lopinavir/Ritonavir (AbbVie, Rungis, France), hydroxychloroquine (Sanofi, Gentilly, France), and others (more than one anti-viral treatment or remsedivir (Gilead, Foster City, USA) as it was not commercially available).

Discrete data were described by their frequency expressed as a percentage together with the 95% confidence interval. Numerical data were described by the mean (with the 95% confidence interval) and standard deviation.

Discrete data were compared using a Chi-square test or Fisher’s exact test, as appropriate. Continuous normally distributed data were compared by ANOVA and continuous non normal data by Kruskal-Wallis as appropriate.

A pre-planned multivariate analysis was performed to identify factors associated with day 28 VFD. We included in the model variables associated with day 28 VFD in univariate analysis with a p value < 0.10 and we forced in the model the type of antiviral strategy. Given 1/ the non-normal distribution of day 28 VFD and 2/ the median value at 0, accordingly to pre-specified rules in the protocol we discriminated the day 28 VFD variables in having at least one day of VFD or not, ie. being extubated and alive at day 28 or not. We performed then a backward conditional logistic regression. Homesher-Lemeshow test and visual inspection were used to ensure the quality of the regression.

A post-hoc multivariate logistic regression on the factors associated with AKI was performed following the same rules given results of univariate analysis. We re-run multivariate logistic regression on the factor associated with the need for renal replacement therapy (RRT).

To account for centre effect, a factor related to patient’s ICU was added in all logistic regression and forced in all models.

No imputation strategy was used for missing data. P value < 0.05 was considered significant. No adjustment was made for multiple comparisons especially for safety outcome [18]. All analyses were performed using Stata (version 16, StataCorp, College Station, TX, USA).

### Ethics Statement

This study was approved by appropriate regulatory committee in France and in Belgium in accordance with national regulation. Each patient was informed about the study. In case of incompetency, next of kin was informed. The requirement for written informed consent was waived due to the study design (France) and after Ethical Committee statement (Belgium).

### Sample size

According to paucity of data on antiviral treatments efficacy, we planned to include at least 250 patients to allow comparison of each antiviral treatment candidate.

## Results

### Baseline characteristics (Table 1)

As of May 10, 380 patients were included in the study and 376 had Day 28 follow up and are reported in the present manuscript. They were mostly male (n=289, 77%), overweighed with mean body mass index (29.8±5.5 kg/m^2^ and suffered from minimal comorbidities previous to COVID-19 as highlighted by Charlson score (1 [0-2]).The most frequent comorbidities is chronic hypertension (n=217 patients; 58%). Patients’ demographic characteristics were similar across different treatment groups (Table 1).

**Table 1:** Patients’ characteristics according to antiviral strategy.

| Total<br>N=376 | No antiviral<br>N= 80 | Lopinavir<br>Ritonavir<br>N= 49 | Hydroxychloroquine<br>N= 197 | Others <sup>a</sup><br>N= 50 | *P Value* |
| --- | --- | --- | --- | --- | --- |
| **Medical History** |  |  |  |  |  |
| Age, mean±SD | 63± 11 | 63 ±11 | 64 ±10 | 62 ±10 | 0.75 |
| Gender, male, n (%) | 59 (74) | 39 (80) | 150 (76) | 41 (82) | 0.7 |
| Body mass index, mean±SD<br>N=78/49/186/7/43 | 30 ±5 | 30 ±8 | 30 ±5 | 30 ±5 | 0.92 |
| History of chronic hypertension, n (%) | 50 (63) | 21 (43) | 116 (59) | 30 (60) | 0.15 |
| Charlson score, median [IQR]<br>N=80/49/197 /7/43 | 1 [0-2] | 1 [0-2] | 1 [0-2] | 0 [0-1] | 0.07 |
| **At inclusion** |  |  |  |  |  |
| Duration between symptoms onset and initiation, median [IQR]<br>N= -/49/192/-/35 | NA | 8 [7-11] | 7 [5-10] | 8 [6-10] | 0.2 |
| Macrolids, n(%) | 33 (41) | 41 (84) | 142 (72) | 21 (42) | <0.001 |
| Including Azithromycin, n =237 (%) | 5 (15) | 3 (7) | 98 (69) | 7 (33) | <0.001 |
| Co-infection, n (%) | 3 (4) | 1 (2) | 34 (17) | 8 (16) | 0.002 |
| **Respiratory values** |  |  |  |  |  |
| Tidal volume, (mL/kg of IBW), mean±SD<br>N=77/49/185/7/43 | 6.2±0.7 | 6.1±0.6 | 6.4±1 | 6.3±0.9 | 0.006 |
| Total PEEP (cmH2O), mean ±SD<br>N=80/49/196/7/43 | 11±3 | 11±3 | 12±3 | 12±3 | 0.04 |
| Plateau pressure (cmH2O), mean ±SD<br>N=65/37/184/7/38 | 23±4 | 22±4 | 24±4 | 23±4 | 0.02 |
| P/F ratio, mean±SD<br>N=80/49/196/7/41 | 131±38 | 146±54 | 124±54 | 117±45 | 0.02 |
| Driving pressure, mean±SD<br>N=64/35/179/-/35 | 13±4 | 12±4 | 13±4 | 11±3 | 0.02 |
| **ARDS treatments** |  |  |  |  |  |
| Neuromuscular blockade, n (%) | 74 (93) | 43 (88) | 156 (79) | 36 (84) | 0.04 |
| Inhaled nitric oxide, n(%) | 4 (5) | 3 (6) | 33 (17) | 3 (6) | 0.01 |
| Prone position, n (%) | 57 (71) | 33 (67) | 169 (86) | 41 (82) | 0.005 |
| **Immunomodulation** |  |  |  |  |  |
| Corticosteroids, n (%)<br>N=354<sup>b</sup> | 10 (16) | 9 (19) | 45 (23) | 13 (26) | 0.6 |
| Inhibitor of IL6, n (%) | 1 (1) | 0 | 8 (4) | 0 | 0.16 |

ARDS: acute respiratory distress syndrome, IBW: ideal body weight, P/F ratio: PaO<sub>2</sub>/FiO<sub>2</sub> ratio.
<sup>a</sup> Bithrapy including HCQ + L/R N= 32; HCQ + Remdesivir N= 11; Remdesivir N=7
<sup>b</sup> some patients were included in a double blind RCT steroids vs placebo (NCT02517489)

The most frequent antiviral treatment administered was hydroxychloroquine monotherapy (HCQ) (n=197; 52%), followed by lopinavir/ritonavir monotherapy (L/R) (n=49; 13%). Other therapy (OTH) (combination or REM) was used in 50 patients (7 REM, 32 HCQ + L/R, 11 HCQ + REM). Of note 80 patients did not receive any supposedly active molecules against SARS-CoV2 (Standard of care, SOC). Antiviral treatments were initiated after 8 [5-10] days of symptoms onset without difference between L/R, HCQ, REM or OTH groups (P=0.2).

### Primary outcomes (Table 2 and Figure 1)

Overall, the number of day 28 VFD was low with a median at 0 [0-13] but significantly different across antiviral strategies (P=0.01). A multivariate logistic regression showed that age (OR 0.95 CI95%: 0.93-0.98), male sex (OR 0.53 CI95% 0.31-0.93) Charlson score (OR 0.85 CI95% 0.73-0.99) and plateau pressure (OR 0.94 CI95% 0.88-0.99) were negatively associated whereas P/F ratio (OR 1.005 CI95% 1.001-1.010) was positively associated with being alive and extubated at day 28 (d28 VFD ≥1). Center effect was not significantly associated with primary outcome (P>0.05). Compared to the SOC strategy, none of the antiviral strategies appeared superior.

**Table 2:** Outcome according to anti-viral treatment.

|  | No antiviral<br>N= 80 | Lopinavir<br>Ritonavir<br>N= 49 | Hydroxychloroquine<br>N= 197 | Others <sup>a</sup><br>N= 50 | *P value* |
| --- | --- | --- | --- | --- | --- |
| **Primary Outcome** |  |  |  |  |  |
| Alive at day 28, n (%) | 58 (73) | 29 (59) | 119 (60) | 36 (72) | 0.14 |
| VFD at day 28, median [IQR]<br>N= 80/49/197/7/43 | 7 [0-15] | 0 [0-11] | 0 [0-12] | 0 [0-9] | 0.01 |
| **Secondary Outcome** |  |  |  |  |  |
| Alive at day 14, n (%) | 65 (81) | 38 (78) | 140 (71) | 41 (82) | 0.2 |
| Ventilatory mode at day 14, n (%) |  |  |  |  | 0.12 |
| - Death | 15 (19) | 11 (22) | 57 (29) | 9 (18) |  |
| - Controlled mode or under VV-ECMO | 24 (30) | 19 (39) | 63 (32) | 21 (42) |  |
| - Pressure Support | 14 (18) | 8 (16) | 38 (19) | 13 (26) |  |
| - Extubated | 27 (34) | 11 (22) | 38 (19) | 7 (14) |  |
| **Safety outcome** |  |  |  |  |  |
| Acute kidney injury, n (%) |  |  |  |  |  |
| - No | 35 (44) | 16 (33) | 94 (48) | 20 (40) | 0.005 |
| - Yes without RRT | 32 (40) | 14 (29) | 65 (33) | 25 (50) |  |
| - Yes with RRT | 13 (16) | 19 (39) | 37 (19) | 5 (10) |  |
| Peak of creatinine, median [IQR]<br>N=79/44/196/7/43 | 117 [84-210] | 188 [91-449] | 122 [80-327] | 147 [90-255] | 0.11 |
| Acute cardiac injury, n (%) | 7 (9) | 5 (10) | 33 (17) | 3 (6) | 0.09 |
| Pulmonary embolism, n (%) | 11 (14) | 4 (8) | 30 (15) | 10 (20) | 0.4 |
| Deep veinous thrombosis, n (%) | 6 (8) | 8 (16) | 17 (9) | 4 (8) | 0.3 |

RRT: Renal replacement therapy, VFD: ventilator free days, VV-ECMO : veno-venous extracorporeal membrane oxygenation
<sup>a</sup> Other including HCQ + L/R N= 32; HCQ + REM N= 11; REM N=7

**Fig. 1.**
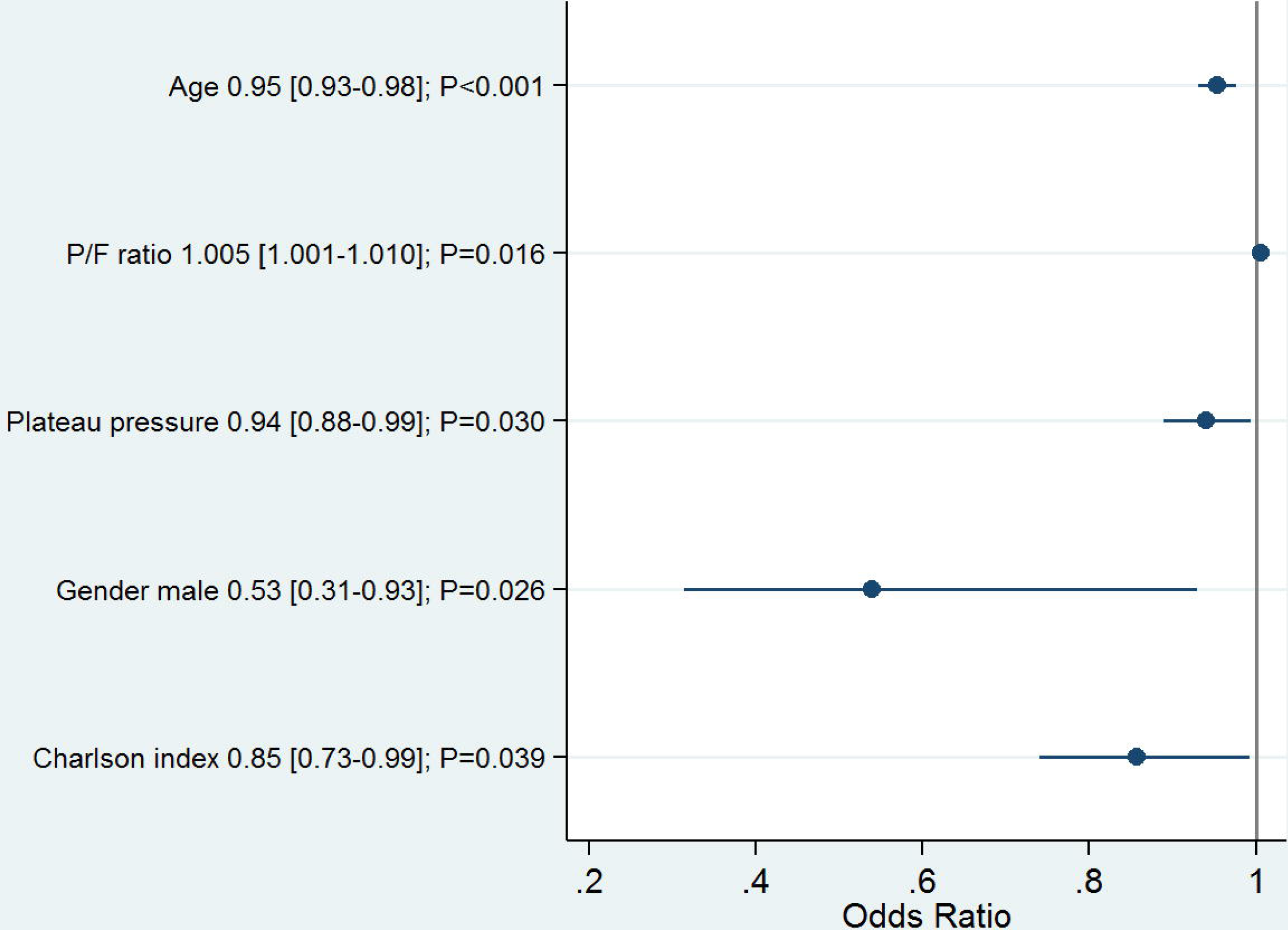
Forrest plot of factors associated with at least one Day 28-VFD in backward multivariate logistic regression (N=328)

### Secondary outcomes (Table 2)

Day 14, and Day 28 survival was similar across treatment groups. At day 14, the distribution of ventilatory modes was similar across treatment groups most of the patients being under controlled mode (Table 2). AKI, Pulmonary Embolism, cardiac injury involved respectively 64%, 15% and 13% of the patients. We observed a difference in AKI repartition in the different groups (P=0.005), whereas the highest creatinine level until day 28 did not differ significantly (P=0.11).

### Factors associated with AKI occurrence (Table 3, Figure 2, eFigure 1)

We compared patients’ characteristics according to the occurrence of AKI (Table 3). Age, BMI, cardiovascular comorbidities, complicated diabetes mellitus and previous chronic kidney diseases were more frequent in patients with AKI. Although, 52 (64%) of the 81 patients that received L/R (either in mono or combination therapy) presented AKI, use of L/R was not significantly more frequent in patients who developed AKI (25% vs 18%; P=0.09). In a post-hoc logistic multivariate regression we identified age, male as gender, history of chronic hypertension, moderate to severe CKD, peripheral arterial disease and use of lopinavir/ritonavir (OR 2.07 CI95% 1.17-3.66) as independent factors associated with AKI (Figure 2). In addition we observed that lopinavir/ritonavir treatment was more frequent in patients that required RRT during their ICU stay (23/73, 31%) as compared with patients without RRT (58/302, 19%; P=0.03). Post-hoc sensitivity logistic regression on the need for RRT provided similar results (eFigure 1). Center effect was not significantly associated with occurrence of AKI nor with need for RRT (P>0.05).

**Table 3:** Associated factors with AKI within 28 days after intubation.

| N=375 | AKI +<br>N=210 | AKI –<br>N=165 | *P value*<sup>1</sup> |
| --- | --- | --- | --- |
| Age, mean±SD | 66 ±9 | 61 ±11) | <0.001 |
| Gender, men, n (%) | 168 (80) | 120 (73) | 0.10 |
| BMI, kg/m<sup>2</sup>, mean±SD | 30.6±6 | 28.9±5 | 0.004 |
| Hypertension, n (%) | 142 (68) | 74 (45) | <0.001 |
| Uncomplicated diabetes mellitus, n (%) | 38 (18) | 31 (19) | 0.9 |
| Complicated diabetes mellitus, n (%) | 24 (11) | 7 (4) | 0.01 |
| Chronic kidney disease, n (%) | 26 (12) | 5 (3) | 0.001 |
| Peripheral artery disease, n (%) | 21 (10) | 3 (2) | 0.001 |
| History of myocardial infarction, n (%) | 26 (12) | 10 (6) | 0.05 |
| Charlson, median [IQR] | 1 (0-3) | 1 (0-1) | <0.001 |
| PEEP (cmH<sub>2</sub>O), mean±SD | 11.7±3 | 11.4±3 | 0.3 |
| Plateau pressure (cmH<sub>2</sub>O), mean±SD<br>N=144/186 | 23.5±4 | 23.6±4 | 0.8 |
| P/F, mean±SD | 126±48 | 129±54 | 0.6 |
| NO, n (%) | 23 (11) | 20 (12) | 0.7 |
| Hydroxychloroquine, n (%) | 126 (60) | 112 (68) | 0.13 |
| Lopinavir/ritonavir, n (%) | 52 (25) | 29 (18) | 0.09 |
| Remdesivir, n (%) | 11 (5) | 7 (4) | 0.8 |
| Corticosteroids<sup>1</sup>, n (%)<br>N=200/153 | 45 (23) | 32 (21) | 0.8 |
| Macrolides | 129 (61) | 107 (65) | 0.5 |
| Co-infection | 28 (13) | 18 (11) | 0.5 |
| Day 14 Ventilatory mode<br>Death<br>Controlled or VV-ECMO<br>Pressure support<br>Extubated | 71 (34)<br>69 (33)<br>40 (19)<br>30 (14) | 21 (13)<br>58 (35)<br>33 (20)<br>53 (32) | < 0.001 |
| Alive at D14, n (%) | 139 (66) | 144 (87) | <0.001 |
| Peak Creatinine until Day 28, median [IQR]<br>n= 205/164 | 267 [148-449] | 80 [64-95] | <0.001 |
| Cardiac injury, n (%) | 40 (19) | 8 (5) | <0.001 |
| Alive at D28, n (%)<br>n=99/88 | 112 (53) | 129 (78) | <0.001 |
| Day 28 VFD, median [IQR]<br>n= 87/81 | 0 (0-2) | 9 (0-17) | < 0.001 |

AKI: acute kidney injury was defined as at least a rise of at least 1.5 times in serum creatinine
BMI: body mass index; SD: standard deviation; P/F: PaO<sub>2</sub>/FiO<sub>2</sub> ratio; NO: Inhaled nitric oxide; VV-ECMO: veno-venous extracorporeal mobile assistance; VFD: ventilator free days
<sup>1</sup> P-value was calculated by Fisher exact test, t-test or Mann Whitney test as appropriate
<sup>2</sup> some patients were included in a RCT steroids vs placebo

**Fig. 2.**
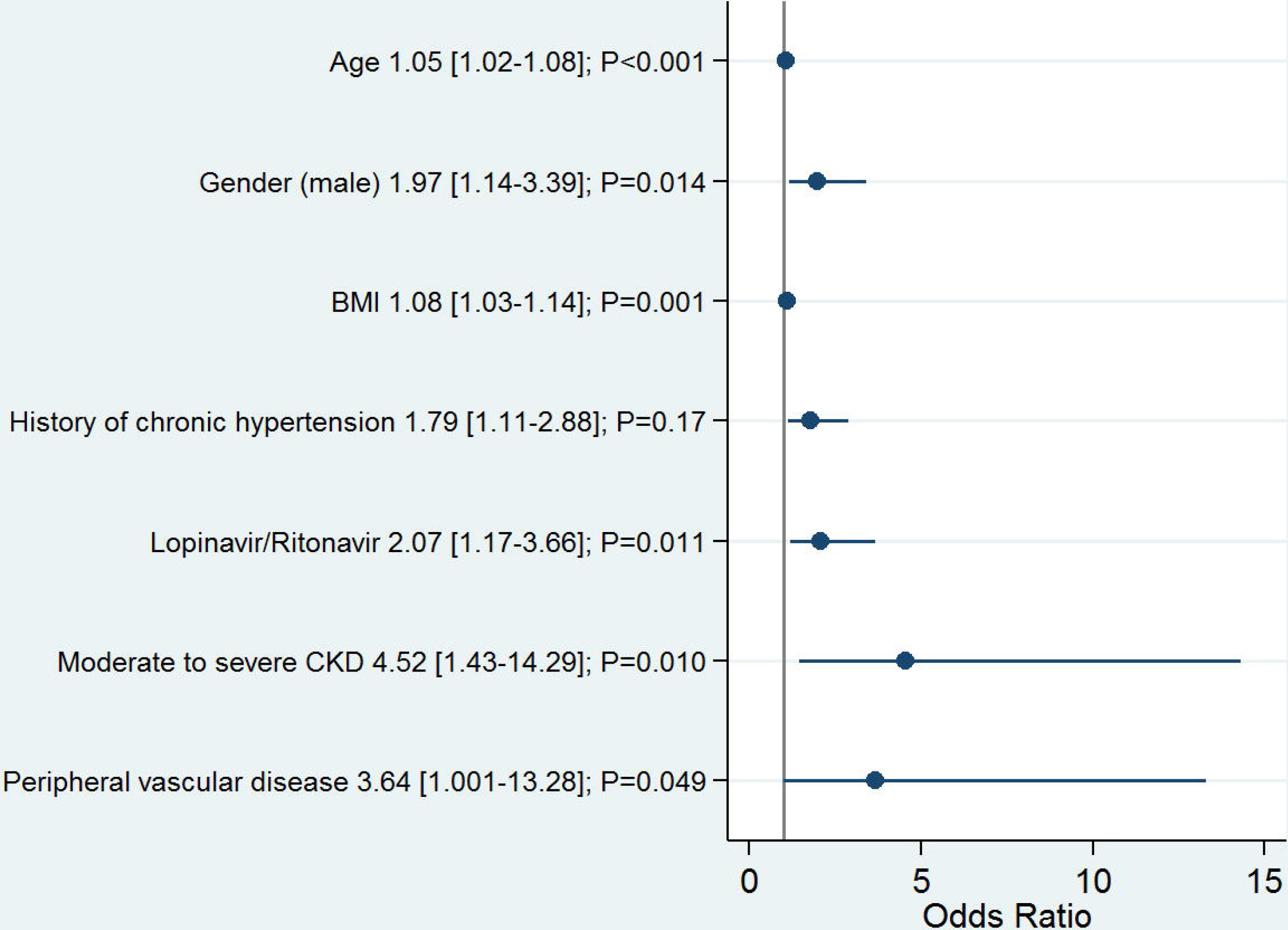
Forrest plot of factors associated with AKI occurrence in backward multivariate logistic regression (N=360)

## Discussion

In this observational study of moderate to severe ARDS complicating COVID-19 in France and Belgium, we did not observe a benefit of treating patients with any of the antiviral molecule under evaluation. However, there was an association between L/R treatment and AKI, including the need for RRT.

Our study is in line with previous findings regarding COVID-19 related ARDS in other countries [6, 19]. Patients were mostly overweighed males between 50 and 70 years of age, with mostly mild cardiovascular comorbidities. Duration of ventilation was high in our cohort exceeding by far the usual duration in ARDS patients (8 [4-16] days in the LUNG-SAFE cohort [20], with day 28 VFD = 10 [0-22]), although day 28 mortality was 36% in our cohort as compared to the 37% in moderate to severe ARDS in the LUNG-SAFE cohort.

Each center managed the ventilator support settings on his own. We observed that in line with ARDS guidelines [21], physicians set tidal volume near 6 mL/kg of IBW, PEEP at moderate-high level, used largely prone positioning and paralysis. We observed significant differences in PEEP level, tidal volume and plateau pressure across patients’ groups (possibly favored by the limited dispersion of the value), but the clinical significance of these differences are uncertain. Hence, although patients were not randomized, we think that the absence of association between any antiviral treatment and positive outcome (neither at day 14 nor at day 28) deserves consideration. Conversely we observed a higher day 28 VFD in the patients without antiviral treatment that questions treatment safety. This finding was not confirmed in the multivariate analysis, which did not show a significant association between any antiviral strategies and day28 VFD. Duration between antiviral initiation and onset of symptoms was in accordance with previous studies: 13 [11-16] days in study by Cao *et al* [7]. While the present study was ongoing, several scientific data emerged on the absence of efficacy of antiviral candidates: for HCQ with possible higher rate of cardiac events especially if combined with azithromycin [22, 23], for L/R in a small randomized controlled trial (RCT) none dedicated to critically ill patients [7]. In parallel, results of US trial dedicated to REM did show a potential benefit on time to recovery but only for patients outside ICU with a mild disease severity [10]. As clinical benefit seems improbable for HCQ, we believe that only the results of ongoing RCTs will determine if REM or L/R have a true effect on outcome, yet we are of the opinion that our data plead to avoid compassionate use of these drugs, following the “first do not harm” rule [24, 25].

Indeed, regarding potential side effects as secondary outcomes, we observed that patients treated by L/R had a higher frequency of renal failure and need for renal replacement therapy. This effect was confirmed after adjustment for potential cofounders associated with a greater risk of renal failure. Several reports in HIV patients indicate that L/R use was associated with an increased risk of chronic kidney disease [26] and acute tubular injury has been described with ritonavir [27]. Renal disease characterized by a proximal tubulopathy have been reported in COVID-19 [28] and have been described as a prognostic factor [29]. We report a high frequency of AKI in our cohort, in line with US data, with 20% of RRT [19]. We observed in our cohort a strong association between day 28-mortality and AKI although we cannot determine if AKI is a marker of a more severe, systemic viral sepsis or a causal determinant of survival but consistent with previous report [30]. To our best knowledge no study dedicated to patient in the ICU analyzed the factors associated with COVID-19 AKI. In our cohort, we observed that besides treatment by L/R, age, gender, BMI, renal and cardiovascular comorbidities were independently associated with AKI. Higher incidence of AKI may then be due either to a previous nephron loss and/or to a specific susceptibility of these patients to SARS-CoV2 through a greater expression of ACE-2 in podocytes and proximal straight tubule cells [31]. Another possible hypothesis is occurrence of hypovolemia related to L/R associated diarrhea and “dry lung” strategy promoted for patients with severe ARDS although usual hemodynamic monitoring in participating ICUs did not support this hypothesis. Last, recent report highlights extremely high dosage for patient with COVID-19 receiving L/R as compare to cohort of HIV-infected patients. [32] Our study was not designed to favor one of these speculative hypotheses. Finally, neither PEEP nor plateau pressure were associated with AKI in opposition with some hypotheses [31].

Finally, we highlight the limitations of our observational study: the patients were not randomized so that we cannot exclude indication bias although collected variables suggest high similarity across treatment strategies. Non-measured confusion biases may exist as well. We did not collect severity score but these scores have been done to compare patients with different diseases in the ICU, and Charlson score, associated with gender and age, have been shown to predict mortality with good accuracy [33]. We have also some missing data which can impact our results. For reasons of lack of time during COVID-19 crisis, we limited strongly the numbers of collected variables so that we are not able to report important data such as the use of ACE inhibitors, or daily ventilator settings. But clinical significance of those factors is also a matter of debate [34]. Lastly, some of these patients have been included in other studies.

## Conclusion

In moderate to severe ARDS COVID-19 patients, we did not observe an association between treatment with hydroxychloroquine or lopinavir/ritonavir and ventilatory free days as compared to no antiviral treatment. We observed an association between lopinavir/ritonavir and AKI. Our data does not support use of these drugs until RCTs results dedicated to patients hospitalized in intensive care will be available.

## Data Availability

The database of the study will be freely accessible online within 3 months after publication upon reasonable request to corresponding author.

## Declarations

### Ethics approval

This study was approved by appropriate regulatory committee in France (CNIL 2217488) and in Belgium (EC n°P2020/253) in accordance with national regulation. Each patient was informed about the study. In case of incompetency, next of kin was informed. The requirement for written informed consent was waived.

### Consent for publication

Not applicable.

## Authors contributions

DG and JBL were responsible for the study concept and design; All authors: acquisition of the data;

DG, JBL, NA, SG, CV, JT: analysis and interpretation of the data; DG and JBL: drafting of the manuscript;

All authors: critical revision of the manuscript for important intellectual content. All authors read and approved the final manuscript.

The corresponding author had full access to all the data in the study and final responsibility for the decision to submit for publication.

## Acknowledgments

We thank Mariana Ismael for Castor EDC (Amsterdam, The Netherlands) for technical support to design eCRF. We thank COVADIS study group investigators:

- Patrick Biston, Intensive Care. CHU-Charleroi, Marie Curie. Université Libre de Bruxelles. 140, Chaussée de Bruxelles. 6042-Charleroi, Belgium
- Gwenhael Colin, Medecine Intensive Reanimation, CHD Vendée, site de la Roche sur Yon, Les Oudairies, 85000 La Roche Sur Yon, France
- Oriane de Maere, Department of Intensive Care,CHR Mons-Hainaut, Mons, Belgium
- Nathan Ebstein, Réanimation médico-chirurgicale CHU Avicennes, Université Sorbonne Paris Nord, Bobigny, France
- Stephan Ehrmann, Médecine Intensive Réanimation, CHRU Tours, Tours, France
- Frederic Foret, Unité de soins intensifs, CHU Dinant Godinne, site Dinant, Belgium
- Lionel Haentjens, Unités de sois intensifs CHU Ambroise Paré, Mons, Belgium
- Thibault Helbert, Réanimation polyvalente Centre Hospitalier du pays d’Aix, Aix en Provence, France
- Jean-Baptiste Mesland, Department of Intensive Care, centres hospitaliers de Jolimont, La Louvière, Belgium
- Celine Monard, Service de réanimation, Hospices Civils de Lyon, 5 Place D’Arsonval, Lyon, France
- Nicolas Mongardon, Service d’anesthésie-réanimation chirurgicale Unité de réanimation chirurgicale polyvalente Hôpitaux Universitaires Henri Mondor, Créteil, France
- Gregoire Ottavy, Medecine Intensive Reanimation, CHU Nantes, 30 Boulevard Jean Monnet, 44093 Nantes Cedex 9, France
- Thomas Pasau, CHU UCL Namur, site Godinne, Av. Dr G. Therasse 1 5530, Yvoir, Belgium
- Gael Piton, Médecine Intensive Réanimation, CHU Besançon, 3 Boulevard FLEMING, 25030 Besançon, France
- Ester Ponzetto, Unité de soins intensifs, Clinique Saint Pierre, Ottignies, Belgium
- Caroline Sejourne, Service de Médecine Intensive Réanimation, CH Germon et Gauthier, Béthune, France
- Morgane Snacken, Soins Intensifs, Hôpital Erasme, ULB, Route de Lennik 808, 1070 Bruxelles, Belgium
- Xavier Souloy, Réanimation - Médecine Intensive, Centre Hospitalier Public du Cotentin, BP208, 50102 Cherbourg-en-Cotentin, France
- Aude Sylvestre, Médecine Intensive Réanimation, Assistance Publique - Hôpitaux de Marseille, Hôpital Nord,, 13015, Marseille, France
- Nicolas Tartrat, Groupe des anesthésistes réanimateurs, Hôpital Privé d’Antony, Antony, France
- Cedric Vanbrussel, Unité de soins intensifs, Clinique Notre Dame de Grâce, Gosselies, Belgium

